# Investigating the integration of sustainable food initiatives in healthcare institutions in Ontario, Canada: A grey literature scoping review protocol

**DOI:** 10.1101/2024.09.28.24314378

**Authors:** Lisa L. Blank, Alyssa D. Milano, Lesley Andrade, Sharon I. Kirkpatrick

## Abstract

**Introduction:** The climate emergency and other sustainability challenges interact to threaten human and planetary health. Efforts to improve the sustainability of food initiatives within healthcare institutions could mitigate these threats by addressing the four pillars of sustainability: health, social, economic, and environmental. Understanding current initiatives to incorporate sustainability into food programs and the sustainability pillars that guide those initiatives is important to inform priorities for action.

This scoping review was undertaken to investigate the extent to which major healthcare institutions in Ontario, Canada have publicly committed to, discussed, planned, and/or implemented sustainable food initiatives.

**Methods and Analysis:** Steps are based on guidance from the Joanna Briggs Institute and Arksey & O’Malley. First, the current strategic plans of 57 healthcare institutions in Ontario, Canada, will be retrieved from their websites and used to examine whether they include any commitments to or discussion, planning, and/or implementation of relevant initiatives. The healthcare institution websites, along with those of selected sustainability organizations, will be searched for grey literature from 2015 to 2024 describing sustainable food initiatives within these institutions. Documents will be screened for eligibility by two researchers. Data related to the incorporation of sustainable food into institutional food programs, and the sustainability pillars addressed, will be extracted by one researcher, with 10% of entries verified by a second researcher. The data will be synthesized to summarize publicly reported progress toward integrating sustainable food into healthcare institutions.

**Ethics and Dissemination:** This review will use publicly available grey literature with no expectation of privacy and no research participants; therefore, no ethics clearance is required.

Results will be shared with stakeholders in sustainability organizations, as well as at relevant conferences and in peer-reviewed journals, such as the Healthy Cities Conference and the Journal of Canadian Food Studies.

This protocol is registered on the Open Science Framework and can be accessed at the following URL: https://doi.org/10.17605/OSF.IO/CU9P6

## Introduction

### Background and Rationale

The climate emergency threatens human and planetary health and the systems we depend on for protection (1,2). Concurrent social, economic, health, and environmental challenges, represented by the Sustainable Development Goals (3), interact with the climate emergency to create significant threats to public health. Food and healthcare are also implicated as drivers and potential responses within this complex socio-ecological system (4–6).

Under the 2015 Paris Agreement, 196 countries committed to enact policies to limit global warming to 2°C above pre-industrial levels while striving to limit warming to 1.5°C (7). In 2023, the Intergovernmental Panel on Climate Change (IPCC) reported that efforts have been grossly inadequate and that if greenhouse gas (GHG) emissions continue at current rates, the world is on track for 2.8°C of warming by 2100 (7). Significant physical and mental health impacts from extreme heat, weather events, infectious disease spread, food insecurity, and infrastructure damage are predicted (2). Climate modelling suggests the social costs of warming will be felt most acutely in human mortality and agricultural losses (8). These effects will interact with the consequences of disturbing many of the planet’s other systems (or planetary boundaries) (9). The modern food system is the primary disruptor of many planetary boundaries including climate change, land-system change, freshwater use, biodiversity, and nitrogen and phosphorous flows (10–13). The strain on the planet’s resources is predicted to increase by 50-92% above present impacts by 2050 from food production and consumption, largely driven by animal-based products and food waste (14).

Challenges to planetary health are occurring alongside a malnutrition epidemic sustained by the modern food system. Overnutrition, in the form of overweight and obesity, affects more than two billion worldwide (5). Concurrently, undernutrition and micronutrient deficiencies continue in global sub-populations (5). These trends have led to increasing rates of diet-related chronic diseases such as diabetes, cardiovascular disease, and cancer, which are the leading causes of disability globally, surpassing tobacco smoking (10).

Nutrition and ecology experts have called for a food system transformation to alter the determinants of food environments, facilitating healthier, more sustainable dietary patterns and food systems (5,10,15–21). Sustainable diets encompass four pillars (also referred to as principles, dimensions, or categories of sustainability), including health, social, economic, and environmental (10,19,21–23). According to the Food and Agricultural Organization of the United Nations (FAO), sustainable diets have “low environmental impacts… contribute to food and nutrition security and to healthy life for present and future generations… (they are) protective and respectful of biodiversity and ecosystems, culturally acceptable, accessible, economically fair and affordable; nutritionally adequate, safe and healthy; while optimizing natural and human resources” (24, p. 7). This holistic conceptualization considers climate and the environment and also includes equity, agency, externalized costs, agricultural livelihoods, disease risk, and leaving a healthy planet for future generations (19,24).

Healthcare is also connected to sustainability. An estimated 5% of Canadian GHG emissions originate from healthcare (25). In 2021, Canada committed to a global agreement to develop climate-resilient, low-carbon health systems (26,27). Publicly funded institutions, including healthcare, are responsible for contributing to this international commitment. Healthcare institutions also have a moral responsibility to lead transformations to more sustainable ways of life (28–30). Part of the impact of healthcare on planetary systems is through food. Healthcare institutions in Canada serve 250,000 meals per day, most of which are procured from large industrial distributors at a total cost of four billion dollars per year (31). The World Health Organization recommends that healthcare institutions use strategies such as promoting local food production, shifting food menus to limit animal-based products, composting food waste, and creating on-site food gardens using Indigenous plants and seeds when possible (6). Shifting toward practices that integrate these recommendations could contribute toward lessening the climate impact of healthcare, with co-benefits for health, local economies and ecosystems (19,28,32,33).

Initiatives such as the Global Green and Healthy Hospitals Network and the World Resources Institute’s Coolfood Pledge indicate awareness of the negative impacts of current food practices within healthcare for climate and the need for action to improve sustainability (6,34,35). According to systematic and scoping reviews conducted to date, limited peer-reviewed studies have considered such efforts and their impacts (36–43). Many initiatives considered in peer-reviewed research focus on the environmental and economic pillars through strategies such as organic and/or local food procurement, waste audits, recycling, and composting (36–44). These initiatives are typically part of institution-wide efforts, not targeted to food, with much of the progress limited to major healthcare institutions in Europe and the United Kingdom (36–45). Little related research has been undertaken within the Canadian context (46,47), yet understanding current commitments and initiatives toward incorporating sustainability into food programs within healthcare is needed to identify priorities for action. Given the broad nature of this issue and lack of prior synthesis, a scoping review is most appropriate (48).

### Objectives

This grey literature scoping review aims to investigate the extent to which major healthcare institutions in Ontario, Canada, have publicly committed to, discussed, planned, and/or implemented sustainability into food programs. The specific objectives are to describe how many major healthcare institutions in Ontario have made strategic commitments to incorporate sustainability and/or sustainable food into their operations; to identify how many of these have publicly discussed, planned, or implemented sustainable food initiatives; and to elucidate the sustainability pillars and strategies considered and/or applied within such initiatives. Distinguishing between levels of progress, such as discussing versus committing, will inform the extent to which sustainability has been integrated.

## Methods

The scoping review design and reporting were informed by guidance from the Joanna Briggs Institute (49) and Arksey and O’Malley (48). Preliminary searches identified that documents discussing efforts related to sustainability within healthcare institutions were more likely to be posted to websites rather than published in peer-reviewed journals. The review will, therefore, focus on grey literature, with a search strategy informed by Godin et al. (50). Stakeholders will be engaged in the search, which has been identified as an effective strategy for supplementing grey literature search results and integrating knowledge mobilization (48,50). Reporting will follow the Preferred Reporting Items for Systematic reviews and Meta-Analyses extension for Scoping Reviews (PRISMA-ScR) (51).

### Eligibility Criteria

The Joanna Briggs Institute’s population, concept, and context (PCC) framework guided the inclusion and exclusion criteria (Table 1) (49). The population will include institutions classified by the *Ontario Public Hospitals Act* (52) (Appendix I) as Group A—general hospitals granted teaching privileges— and Group B—hospitals with no fewer than 100 beds. Documents describing these institutions will be eligible for inclusion (n=57 institutions) (52). Institutions with fewer than 100 beds and those providing specialized care, such as rehabilitation or psychiatric care, will be excluded (52). Major healthcare institutions with larger budgets and a higher number of beds are hypothesized to have more resources to allocate to sustainability-related initiatives, as well as having a wider reach through larger catchment areas, than smaller institutions. Documents relevant to campuses of parent health corporations with at least 100 beds will be considered. If few documents related to the sustainability of food programs within these 57 institutions are identified, the scope may be expanded within Ontario as well as to other provinces and territories within Canada.

**Table 1.** Eligibility Criteria.

| Inclusion Criteria | Exclusion Criteria |
| --- | --- |
| Related to major general healthcare institutions granted teaching privileges and those with no fewer than 100 beds in Ontario, Canada | Describes initiatives in institutions in Ontario with fewer than 100 beds and those providing specialized care or in institutions outside of Ontario |
| Mentions sustainability in the context of institutional food services, including any of the four pillars: health, social, economic, and/or environmental | Does not mention sustainability in the context of institutional food services |
| Grey literature including reports, press releases, blog posts, newsletters | Peer-reviewed literature<br>Social media posts on Instagram, Facebook, YouTube, LinkedIn, or TikTok |
| Publicly available online | Access controlled by subscription or membership |
| Dated between 2015 and 2024 | Dated before 2015 |

The most recent, publicly available strategic plans of the 57 healthcare institutions, or their parent health organizations, will be eligible for inclusion regardless of whether they mention sustainability or their publication date. Information from these reports will be used to determine the level of progress (commitment, discussion, planning, and/or implementation) toward integrating sustainability into institutional programs.

Beyond strategic plans, documents related to the 57 institutions that include the concept of sustainability within at least one of its four pillars—health, social, economic, and environmental— will be eligible for inclusion. For example, documents describing interventions to reduce diet-related chronic diseases, improve the cultural appropriateness of menus, strengthen local food production, and/or reduce food waste will be within scope. Documents pertaining to sustainability in the context of other operations, such as energy use, green facility construction, or reduction of anesthetic gases, will be excluded.

Grey literature documents may include publicly accessible reports, press releases, blog posts, and newsletters. Documents excluded from consideration will include internal communications (e.g., via institutional intranets) and those behind paywalls because their information is not publicly accessible. Posts to social media will be excluded on the basis that meaningful commitments and/or actions related to sustainability are expected to extend beyond social media posts.

Documents dated between 2015 and 2024 will be considered for inclusion. The Sustainable Development Goals, adopted in September 2015, were landmark commitments to sustainability, integrating food, health, climate, and other planetary systems (3,53). Additionally, in 2015, experts announced that four of the nine planetary boundaries had been crossed (11), and the Paris Agreement was adopted (7). When necessary, the Wayback Machine will be used to identify the publishing date of undated documents (54). Documents available in English or French will be eligible for inclusion. French documents will be translated using DeepL, an online translation application (55).

Any changes to the eligibility criteria and the rationale will be described in full in the scoping review.

### Information Sources and Search Strategy

The most recent strategic plans available on the websites of the 57 institutions, or the websites of the parent organization, will be retrieved. Next, the healthcare institutions’ websites will be searched for grey literature documents that specifically discuss sustainability in the context of food, using the search function within each website. The searches will use terms to capture the population, concept, and context (Table 2), with combinations of the search terms applied depending on each website’s search capability. The URLs, search terms, and retrieval dates of the first 25 results for each unique search will be recorded in an Excel (Microsoft Corporation, Redmond, Washington, United States) spreadsheet. Zotero (Corporation for Digital Scholarship, Vienna, Virginia, United States) will be used to manage citations.

**Table 2.** Search Terms.

| Population | Concept | Context |
| --- | --- | --- |
| Hospital | Sustainable food | Ontario, Canada |
| Healthcare institution | Planetary health diet |  |
| Healthcare facility | Eco-friendly nutrition |  |
| Healthcare centre |  |  |

Additionally, the websites of seven organizations involved in sustainability in Canadian healthcare institutions will be searched using each website’s search function. These organizations include Nourish Leadership, Cascades, PEACH Health Ontario, the Canadian Coalition for Green Health Care, HealthcareCAN, Healthcare without Harm, and Practice GreenHealth (26,35,56–60). Searching these websites is intended to identify information related to efforts to integrate sustainability in Ontario healthcare institutions that may not have been located on institutional websites. In preliminary searches, documents relevant to the review objectives were located on these organization’s websites. Combinations of search terms may be used depending on each website’s search capability. The URLs, search terms, and retrieval dates for each unique search will be recorded in the Excel (Microsoft, Redmond, Washington) spreadsheet.

### Screening

Duplicates identified through the searches will be identified using Excel (Microsoft Corporation, Redmond, Washington, United States) and removed. The strategic plans will automatically proceed to data extraction. As grey literature may not include abstracts or executive summaries, the full text of the remaining retrieved documents will be screened by two reviewers based on the eligibility criteria. Pilot screening of 25 documents will be conducted until an agreement rate exceeding 90% is reached. The remainder of the documents will then be screened independently by the two reviewers, with discrepancies discussed with a third reviewer to reach consensus. The screening result and, if applicable, the rationale for exclusion will be recorded. The URLs, search terms, retrieval date, publishing date, institution(s) cited, document type, and whether sustainable food initiatives were discussed will be retained for all documents (Appendix II). A PRISMA flow diagram will summarize all stages of document retrieval, screening, and eligibility for inclusion.

### Data Extraction

An Excel (Microsoft, Redmond, Washington, United States) spreadsheet has been developed to guide data extraction (Appendix II). Data extraction will be piloted with 10% of the relevant documents by two researchers to ensure that the data extraction fields are accurately charted. Following discussion and resolution of any discrepancies, the remaining data extraction will be conducted by one researcher.

For strategic reports, the full documents will be reviewed for discussion of any of the four pillars of sustainability related to any aspect of the institution’s operations, including but not exclusive of food. Descriptions of these commitments to sustainability and any descriptions of further discussion, planning, and/or implementation will be extracted.

The remaining documents will be reviewed for evidence of progress toward strategic commitments, such as discussion, planning, and/or implementation, or an intention to incorporate sustainable food initiatives not outlined in strategic plans. Relevant details related to efforts to incorporate sustainable food, including cited institution(s), department(s) involved, intended outcome(s), and/or associated activities, will be extracted (Appendix II). If applicable, timelines and any efforts to evaluate initiatives will be recorded.

### Stakeholder Consultation

Following the completion of data extraction, the Excel (Microsoft, Redmond, Washington) spreadsheet will be shared via email with stakeholders working to improve sustainability in Canadian healthcare institutions. Preliminary connections have been made with representatives from the sustainability organizations included in the search strategy, who have relevant expertise and experience. Stakeholders will be asked to identify any additional documents relevant to initiatives within the 57 hospitals. Only publicly available documents with no expectation of privacy will be included. Any documents shared will then undergo screening and data extraction according to the procedures above.

### Synthesis and Presentation of Results

Using data extracted from the strategic plans, institutional commitments to incorporate sustainability into overall and food-related operations will be synthesized narratively. Next, using data extracted from the remaining documents, evidence of progress toward commitments and sustainable food initiatives beyond institutional commitments will be synthesized narratively. Evidence of progress will be interpreted based on whether documents describe discussions related to one or more possible relevant initiatives, describe the planning of specific activities towards one or more initiatives, or describe the implementation of one or more initiatives. The 57 healthcare institutions will be classified according to whether they have made strategic commitments to incorporate sustainability into food programs, whether they have made progress toward those commitments, the stage of progress, and/or whether they have created sustainable food initiatives not mentioned in strategic commitments. Figures will illustrate frequency counts.

To understand the nature of current initiatives, the sustainability pillars and strategies used will also be synthesized narratively and using charts. This synthesis will draw on details extracted from retrieved documents, including intended outcome(s), department(s) involved, and associated activities. Relevant timelines and evaluation efforts will also be summarized narratively to further inform the extent of progress. The location of institutions discussing, planning and/or implementing initiatives will be mapped to inform regions of the province with more and less progress. The extent of sustainable food integration will be compared to other provinces in Canada and examples internationally.

Any changes to the planned synthesis and presentation approach will be reported in full in the scoping review.

### Ethics and Dissemination

This research will be a scoping review of publicly available grey literature with no expectation of privacy and no research participants. Therefore, the University of Waterloo’s Research Ethics Board required no ethics approval. Nonetheless, care will be taken not to report identifiable information for stakeholders or persons described in retrieved documents.

## Conclusion

This will be the first grey literature scoping review investigating efforts to integrate sustainable food and describing the most common sustainability pillars and strategies applied within Ontario healthcare institutions. While some information may be missed by relying on digital sources, this review is meant to capture the extent of publicly available progress integrating sustainability by distinguishing between commitments, discussion, planning, and implementation. The aim is to compile a broad summary of current efforts in healthcare institutions in Ontario to inform future endeavours in Ontario, across Canada, and internationally to mitigate and adapt to the climate emergency and other sustainability challenges.

## Supporting information

PRISMA-ScR Checklist

## Data Availability

All data produced in the present study are available upon reasonable request to the authors.

## Acknowledgements

Dr. Kirsten M. Lee contributed to conceptualizing this scoping review in its earliest stages.

## Appendices

### Appendix I: *Ontario Public Hospitals Act* Classification of Hospitals as of May 31, 2024

Group A hospitals: general hospitals providing facilities for giving instruction to medical students and for providing post-graduate education leading to certification or a fellowship

Group B hospitals: having no fewer than 100 beds

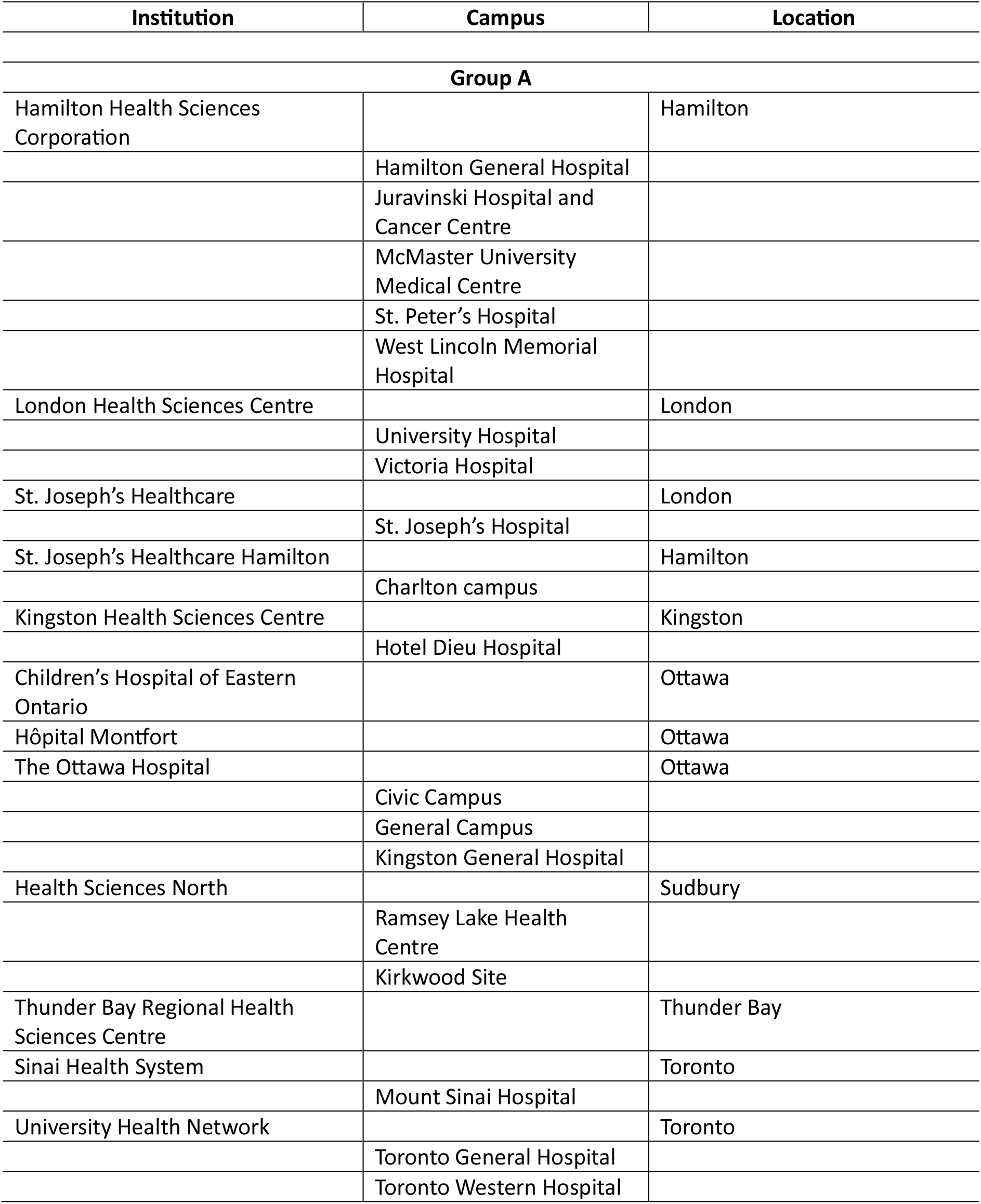
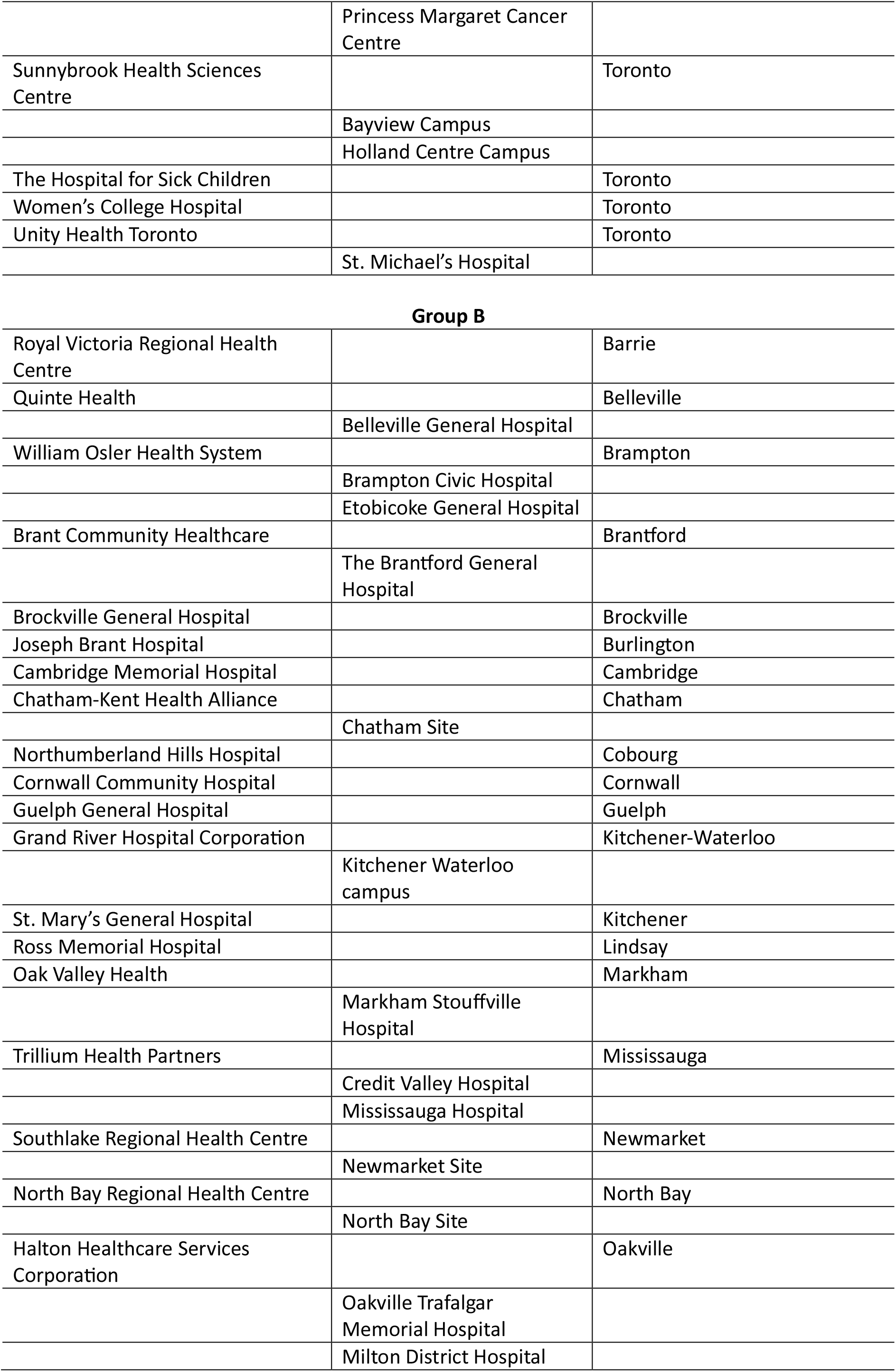
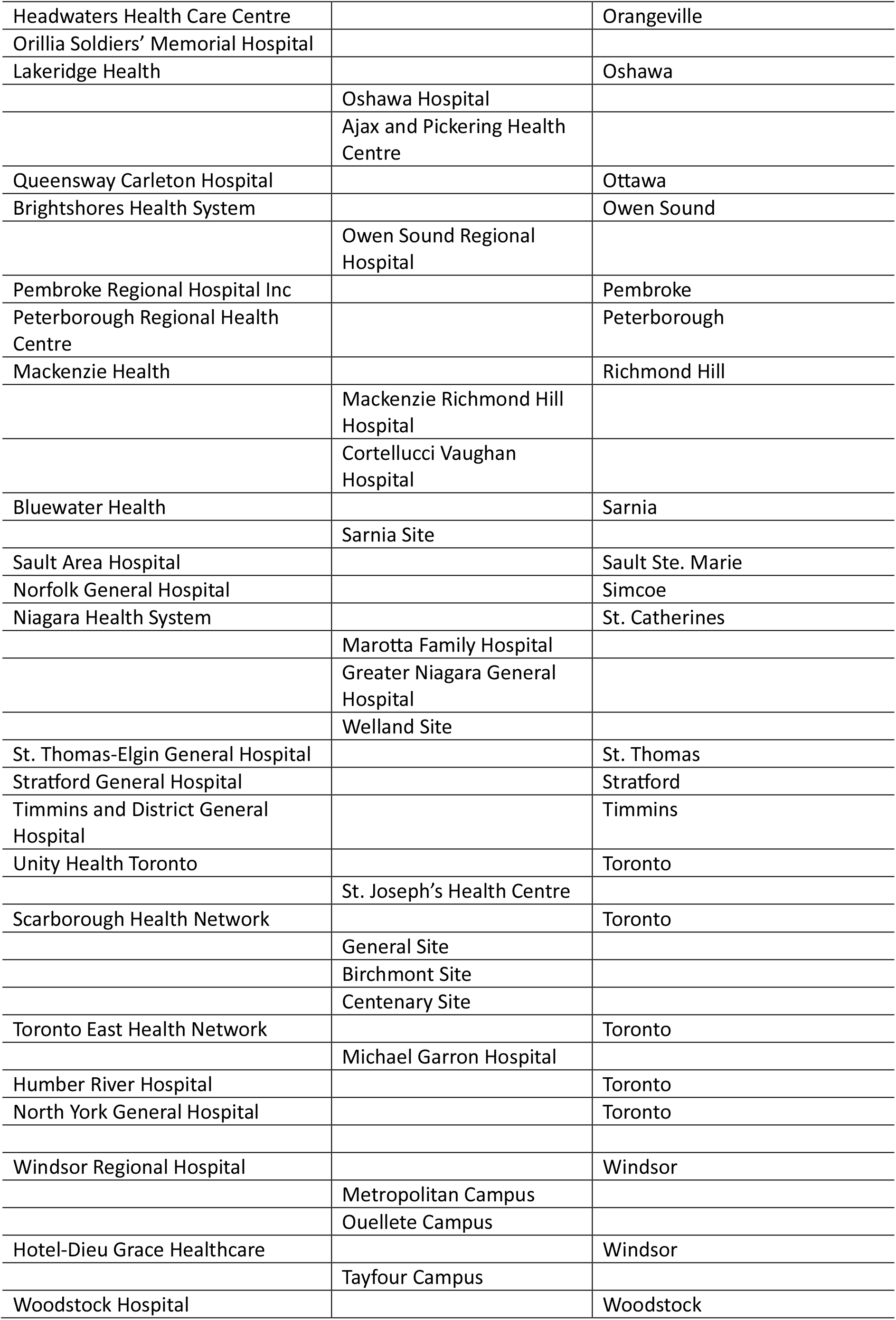

| Institution | Campus | Location |
| --- | --- | --- |
| **Group A** |  |  |
| Hamilton Health Sciences Corporation |  | Hamilton |
|  | Hamilton General Hospital |  |
|  | Juravinski Hospital and Cancer Centre |  |
|  | McMaster University Medical Centre |  |
|  | St. Peter's Hospital |  |
|  | West Lincoln Memorial Hospital |  |
| London Health Sciences Centre |  | London |
|  | University Hospital |  |
|  | Victoria Hospital |  |
| St. Joseph's Healthcare |  | London |
|  | St. Joseph's Hospital |  |
| St. Joseph's Healthcare Hamilton |  | Hamilton |
|  | Charlton campus |  |
| Kingston Health Sciences Centre |  | Kingston |
|  | Hotel Dieu Hospital |  |
| Children's Hospital of Eastern Ontario |  | Ottawa |
| Hôpital Montfort |  | Ottawa |
| The Ottawa Hospital |  | Ottawa |
|  | Civic Campus |  |
|  | General Campus |  |
|  | Kingston General Hospital |  |
| Health Sciences North |  | Sudbury |
|  | Ramsey Lake Health Centre |  |
|  | Kirkwood Site |  |
| Thunder Bay Regional Health Sciences Centre |  | Thunder Bay |
| Sinai Health System |  | Toronto |
|  | Mount Sinai Hospital |  |
| University Health Network |  | Toronto |
|  | Toronto General Hospital |  |
|  | Toronto Western Hospital |  |
|  | Princess Margaret Cancer Centre |  |
| Sunnybrook Health Sciences Centre |  | Toronto |
|  | Bayview Campus |  |
|  | Holland Centre Campus |  |
| The Hospital for Sick Children |  | Toronto |
| Women's College Hospital |  | Toronto |
| Unity Health Toronto |  | Toronto |
|  | St. Michael's Hospital |  |
| Group B |  |  |
| Royal Victoria Regional Health Centre |  | Barrie |
| Quinte Health |  | Belleville |
|  | Belleville General Hospital |  |
| William Osler Health System |  | Brampton |
|  | Brampton Civic Hospital |  |
|  | Etobicoke General Hospital |  |
| Brant Community Healthcare |  | Brantford |
|  | The Brantford General Hospital |  |
| Brockville General Hospital |  | Brockville |
| Joseph Brant Hospital |  | Burlington |
| Cambridge Memorial Hospital |  | Cambridge |
| Chatham-Kent Health Alliance |  | Chatham |
|  | Chatham Site |  |
| Northumberland Hills Hospital |  | Cobourg |
| Cornwall Community Hospital |  | Cornwall |
| Guelph General Hospital |  | Guelph |
| Grand River Hospital Corporation |  | Kitchener-Waterloo |
|  | Kitchener Waterloo campus |  |
| St. Mary's General Hospital |  | Kitchener |
| Ross Memorial Hospital |  | Lindsay |
| Oak Valley Health |  | Markham |
|  | Markham Stouffville Hospital |  |
| Trillium Health Partners |  | Mississauga |
|  | Credit Valley Hospital |  |
|  | Mississauga Hospital |  |
| Southlake Regional Health Centre |  | Newmarket |
|  | Newmarket Site |  |
| North Bay Regional Health Centre |  | North Bay |
|  | North Bay Site |  |
| Halton Healthcare Services Corporation |  | Oakville |
|  | Oakville Trafalgar Memorial Hospital |  |
|  | Milton District Hospital |  |
| Headwaters Health Care Centre |  | Orangeville |
| Orillia Soldiers' Memorial Hospital |  |  |
| Lakeridge Health |  | Oshawa |
|  | Oshawa Hospital |  |
|  | Ajax and Pickering Health Centre |  |
| Queensway Carleton Hospital |  | Ottawa |
| Brightshores Health System |  | Owen Sound |
|  | Owen Sound Regional Hospital |  |
| Pembroke Regional Hospital Inc |  | Pembroke |
| Peterborough Regional Health Centre |  | Peterborough |
| Mackenzie Health |  | Richmond Hill |
|  | Mackenzie Richmond Hill Hospital |  |
|  | Cortellucci Vaughan Hospital |  |
| Bluewater Health |  | Sarnia |
|  | Sarnia Site |  |
| Sault Area Hospital |  | Sault Ste. Marie |
| Norfolk General Hospital |  | Simcoe |
| Niagara Health System |  | St. Catharines |
|  | Marotta Family Hospital |  |
|  | Greater Niagara General Hospital |  |
|  | Welland Site |  |
| St. Thomas-Elgin General Hospital |  | St. Thomas |
| Stratford General Hospital |  | Stratford |
| Timmins and District General Hospital |  | Timmins |
| Unity Health Toronto |  | Toronto |
|  | St. Joseph's Health Centre |  |
| Scarborough Health Network |  | Toronto |
|  | General Site |  |
|  | Birchmont Site |  |
|  | Centenary Site |  |
| Toronto East Health Network |  | Toronto |
|  | Michael Garron Hospital |  |
| Humber River Hospital |  | Toronto |
| North York General Hospital |  | Toronto |
| Windsor Regional Hospital |  | Windsor |
|  | Metropolitan Campus |  |
|  | Ouellete Campus |  |
| Hotel-Dieu Grace Healthcare |  | Windsor |
|  | Tayfour Campus |  |
| Woodstock Hospital |  | Woodstock |

### Appendix II: Data screening and extraction fields from Excel spreadsheet

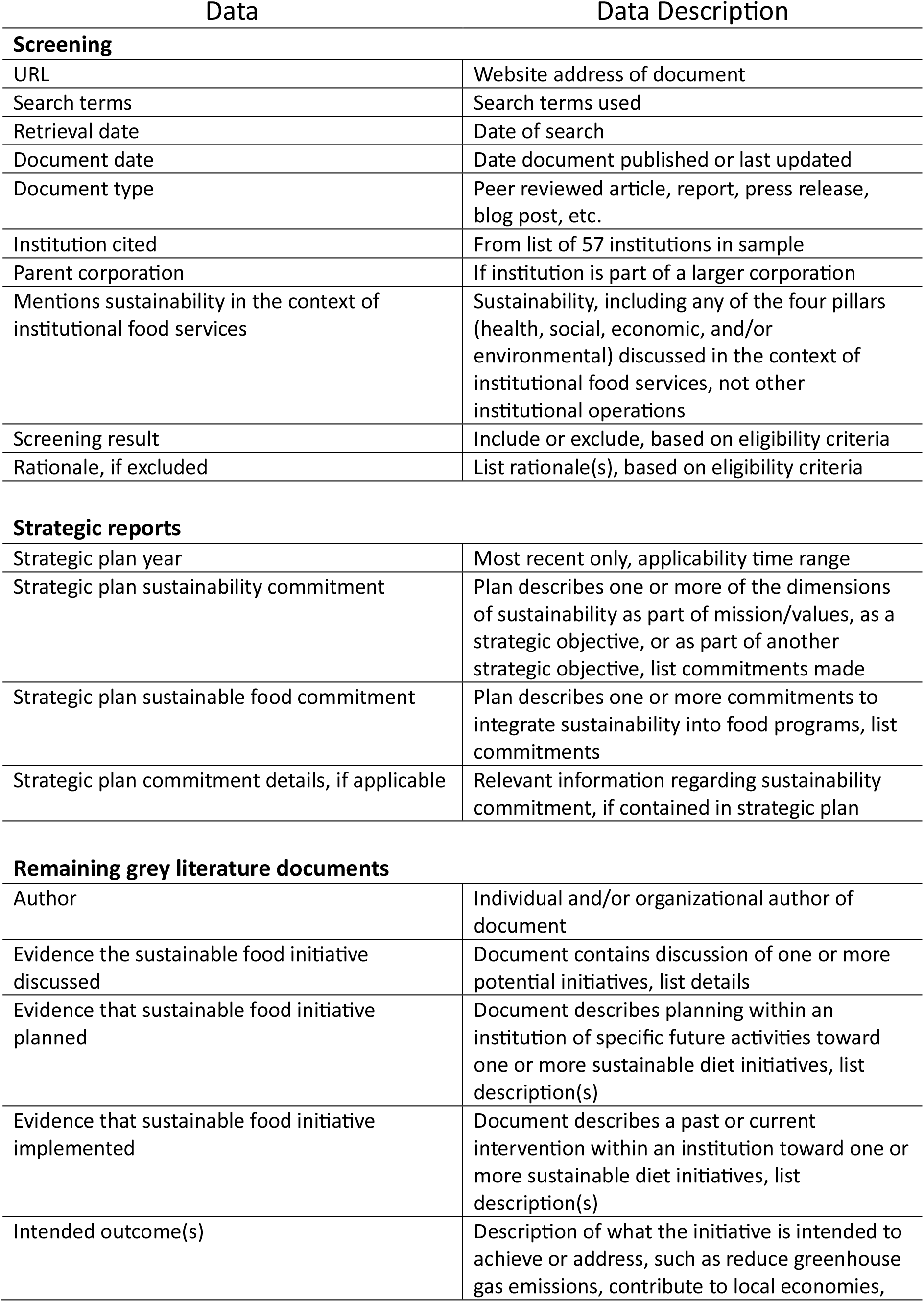
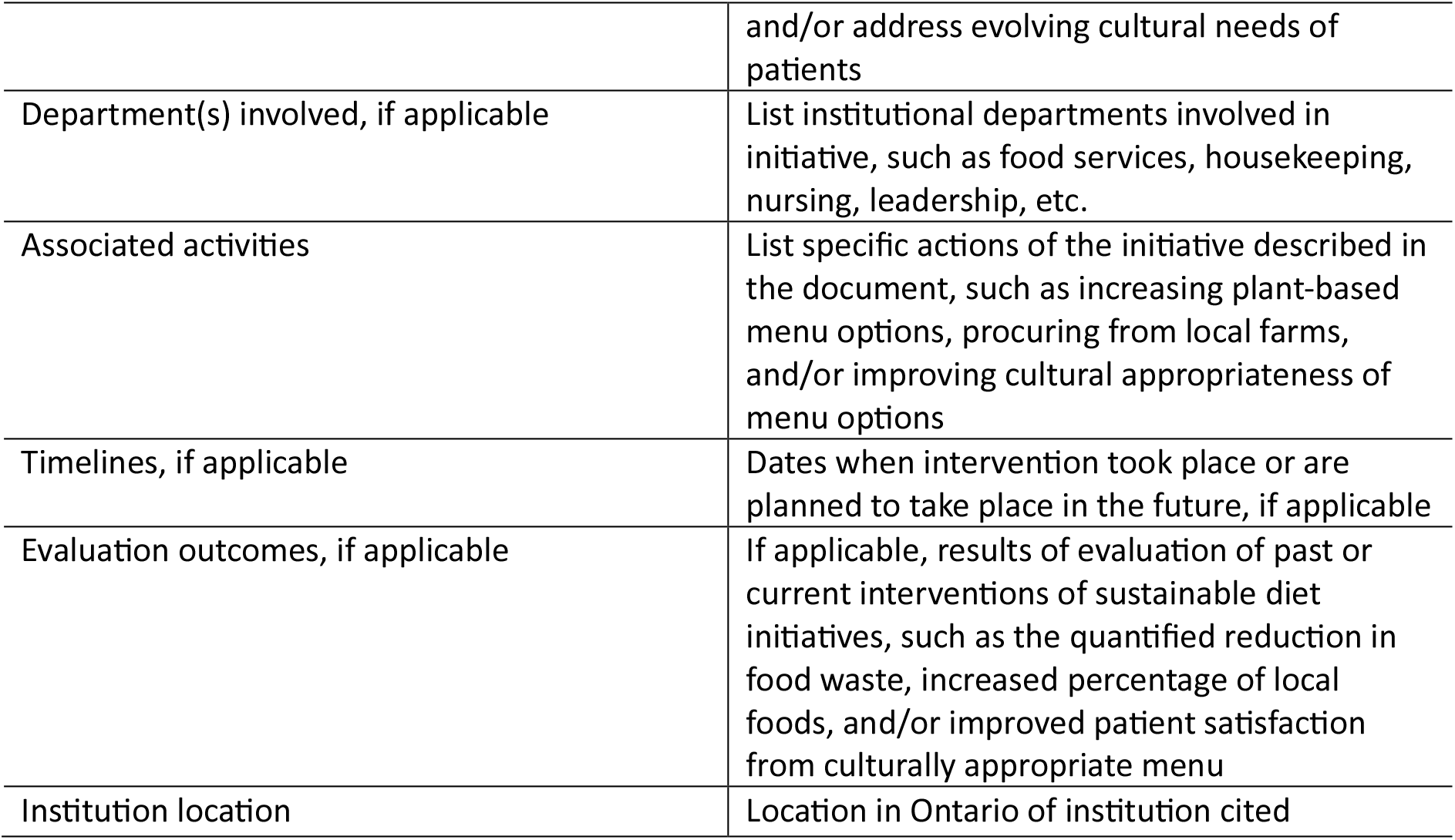

| Data |  | Data Description |
| --- | --- | --- |
| **Screening** |  |  |
| URL |  | Website address of document |
| Search terms |  | Search terms used |
| Retrieval date |  | Date of search |
| Document date |  | Date document published or last updated |
| Document type |  | Peer reviewed article, report, press release, blog post, etc. |
| Institution cited |  | From list of 57 institutions in sample |
| Parent corporation |  | If institution is part of a larger corporation |
| Mentions sustainability in the context of institutional food services |  | Sustainability, including any of the four pillars (health, social, economic, and/or environmental) discussed in the context of institutional food services, not other institutional operations |
| Screening result |  | Include or exclude, based on eligibility criteria |
| Rationale, if excluded |  | List rationale(s), based on eligibility criteria |
| **Strategic reports** |  |  |
| Strategic plan year |  | Most recent only, applicability time range |
| Strategic plan sustainability commitment |  | Plan describes one or more of the dimensions of sustainability as part of mission/values, as a strategic objective, or as part of another strategic objective, list commitments made |
| Strategic plan sustainable food commitment |  | Plan describes one or more commitments to integrate sustainability into food programs, list commitments |
| Strategic plan commitment details, if applicable |  | Relevant information regarding sustainability commitment, if contained in strategic plan |
| **Remaining grey literature documents** |  |  |
| Author |  | Individual and/or organizational author of document |
| Evidence the sustainable food initiative discussed |  | Document contains discussion of one or more potential initiatives, list details |
| Evidence that sustainable food initiative planned |  | Document describes planning within an institution of specific future activities toward one or more sustainable diet initiatives, list description(s) |
| Evidence that sustainable food initiative implemented |  | Document describes a past or current intervention within an institution toward one or more sustainable diet initiatives, list description(s) |
| Intended outcome(s) |  | Description of what the initiative is intended to achieve or address, such as reduce greenhouse gas emissions, contribute to local economies, |

|  |  |
| --- | --- |
|  | and/or address evolving cultural needs of patients |
| Department(s) involved, if applicable | List institutional departments involved in initiative, such as food services, housekeeping, nursing, leadership, etc. |
| Associated activities | List specific actions of the initiative described in the document, such as increasing plant-based menu options, procuring from local farms, and/or improving cultural appropriateness of menu options |
| Timelines, if applicable | Dates when intervention took place or are planned to take place in the future, if applicable |
| Evaluation outcomes, if applicable | If applicable, results of evaluation of past or current interventions of sustainable diet initiatives, such as the quantified reduction in food waste, increased percentage of local foods, and/or improved patient satisfaction from culturally appropriate menu |
| Institution location | Location in Ontario of institution cited |

